# COVID-19 data analysis and modeling in Palestine

**DOI:** 10.1101/2020.04.24.20078279

**Authors:** Ines Abdeljaoued-Tej

## Abstract

We estimate an actual number of infected cases in Palestine based on the 18-day effect from infection to death. We find that the number of cases in April 22 varies between 506 and 2 026 infected cases. We also focus on the reproductive number in Palestine based on population dynamics with two SEIR models. Dataset is from 5 March to 22 April 2020. With a transmission rate equal to 4.55 10^−6^, on May 22, the simulations predict 11 014 total infected cases in the optimistic scenario and 113 171 in the worst one. The crest of the pandemic is from 22 to 27 May 2020. The reproductive number ℛ_0_ is equal to 1.54 for a fixed fraction of 0.6 of symptomatic cases that are reported and for a removal rate of 7. Palestinian COVID-19 mortality number is equal to 6 per million. It is small compared to countries neighboring Palestine. The infected number is equal to 88.4 per million, which is less than most of its neighbors. The basic reproduction number is still greater than 1. Changes to the transmission rate (over time) would be advisable, to fall ℛ_0_ below the critical threshold.

## 1 Introduction

The novel coronavirus SARS-CoV-2 (COVID-19), has spread to many countries. Given the fragile health systems in most countries, and especially in war zones, we studied the data available in Palestine as well as their possible evolution. We address the following fundamental issues concerning this epidemic: How will the epidemic evolve in Palestine concerning the number of reported cases and unreported cases? How will the number of unreported cases influence the severity of the epidemic? Most of the Palestinian territories live under a blockade and apartheid. What is the effect of these oppressive policies on the epidemic? To answer these questions, we developed mathematical models that recover from data of reported cases and the number of unreported cases for the COVID-19 epidemic in Palestine.

We use a set of reported data to model the epidemic in Palestine: data of Table 4 is shared by the Palestinian Ministry of Health. It represents the epidemic transmission in Palestine. The first case was detected on March 5, 2020. Four deaths are reported on 22 April 2020 with more than 466 total number of infected cases. The three phases of COVID-19 epidemics can be decomposed as a linear phase, an exponential growth, and a decreasing stage. The linear growth in the number of reported cases (from 5 March to 31 March) is where the number of daily reported cases is almost constant day after day. The second phase of the epidemic corresponds to an exponential increasing phase, it starts on April 1st, 2020. The third phase of the epidemic corresponds to a time-dependent exponentially decreasing transmission rate, due to major public interventions and social distancing measures [5]. Our analysis identifies the epidemics in Palestine as in the exponential phase. The main objective of this study is the estimation of the average number of infections one case can generate throughout the infectious period. It is the basic reproduction number of an infectious agent. Section 2 presents the data set and an estimation of an actual number of infected cases in Palestine based on the 18-day effect from infection to death.

**Table 4:** Data for Palestine - accumulated reported cases, *CR*(*t*), and casualties

| Date | Infected | Death |
| --- | --- | --- |
| 2020-03-04 | 0 | 0 |
| 2020-03-05 | 7 | 0 |
| 2020-03-06 | 16 | 0 |
| 2020-03-07 | 19 | 0 |
| 2020-03-08 | 19 | 0 |
| 2020-03-09 | 26 | 0 |
| 2020-03-10 | 30 | 0 |
| 2020-03-11 | 30 | 0 |
| 2020-03-12 | 31 | 0 |
| 2020-03-13 | 35 | 0 |
| 2020-03-14 | 38 | 0 |
| 2020-03-15 | 38 | 0 |
| 2020-03-16 | 39 | 0 |
| 2020-03-17 | 44 | 0 |
| 2020-03-18 | 44 | 0 |
| 2020-03-19 | 47 | 0 |
| 2020-03-20 | 48 | 0 |
| 2020-03-21 | 52 | 0 |
| 2020-03-22 | 59 | 0 |
| 2020-03-23 | 59 | 0 |
| 2020-03-24 | 60 | 1 |
| 2020-03-25 | 71 | 1 |
| 2020-03-26 | 84 | 1 |
| 2020-03-27 | 91 | 1 |
| 2020-03-28 | 98 | 1 |
| 2020-03-29 | 108 | 1 |
| 2020-03-30 | 117 | 1 |
| 2020-03-31 | 119 | 1 |
| 2020-04-01 | 133 | 1 |
| 2020-04-02 | 161 | 1 |
| 2020-04-03 | 194 | 1 |
| 2020-04-04 | 217 | 1 |
| 2020-04-05 | 237 | 1 |
| 2020-04-06 | 254 | 1 |
| 2020-04-07 | 261 | 1 |
| 2020-04-08 | 263 | 1 |
| 2020-04-09 | 263 | 2 |
| 2020-04-10 | 267 | 2 |
| 2020-04-11 | 268 | 2 |
| 2020-04-12 | 290 | 2 |
| 2020-04-13 | 308 | 2 |
| 2020-04-14 | 374 | 2 |
| 2020-04-15 | 374 | 2 |
| 2020-04-16 | 402 | 2 |
| 2020-04-17 | 418 | 2 |
| 2020-04-18 | 439 | 3 |
| 2020-04-19 | 449 | 3 |

Data information includes the cumulative number of reported cases, as shown in Figure 1. Section 3 introduces two deterministic compartmental model based on the clinical progression of the disease and the epidemiological status of the individuals [7]. The SEIR model studies the disease transmission. Four compartments are present: Susceptible, Exposed, Infectious, and Recovered. The cumulative number of reported symptomatic infectious cases at time *t*, denoted by *CR*(*t*) is computed. We then construct numerical simulations and compare them with data.

**Figure 1:**
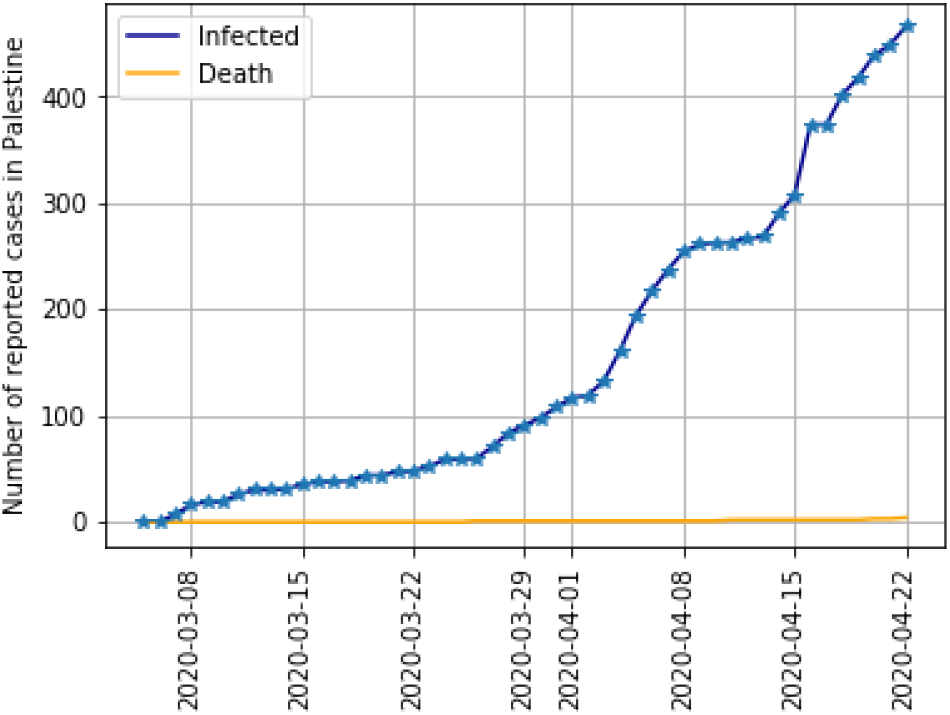
Reported cases in Palestine until 22^*th*^ April 2020. The number of deaths is 4. This amount represents 6.68‰ of total cases.

## 2 Data description

The data available in [2] gives the number of COVID-19 positive cases per day, the number of deaths due to COVID-19, and the number of reported recoveries. But this repository does not contain data from Palestine. The data entry was done manually^1^. The daily growth rate of the infected cases in Figure 2 is given by: 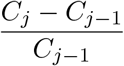 in day *j*, where *C*_*j*_ is the number of reported COVID-19 cases at time *j*. The median of the daily growth rate of reported cases in Palestine from March 6 to 22^*th*^ April 2020 is equal to 0.07. The total number of deaths is 4. It represents 6.7 ‰ of total reported cases.

**Figure 2:**
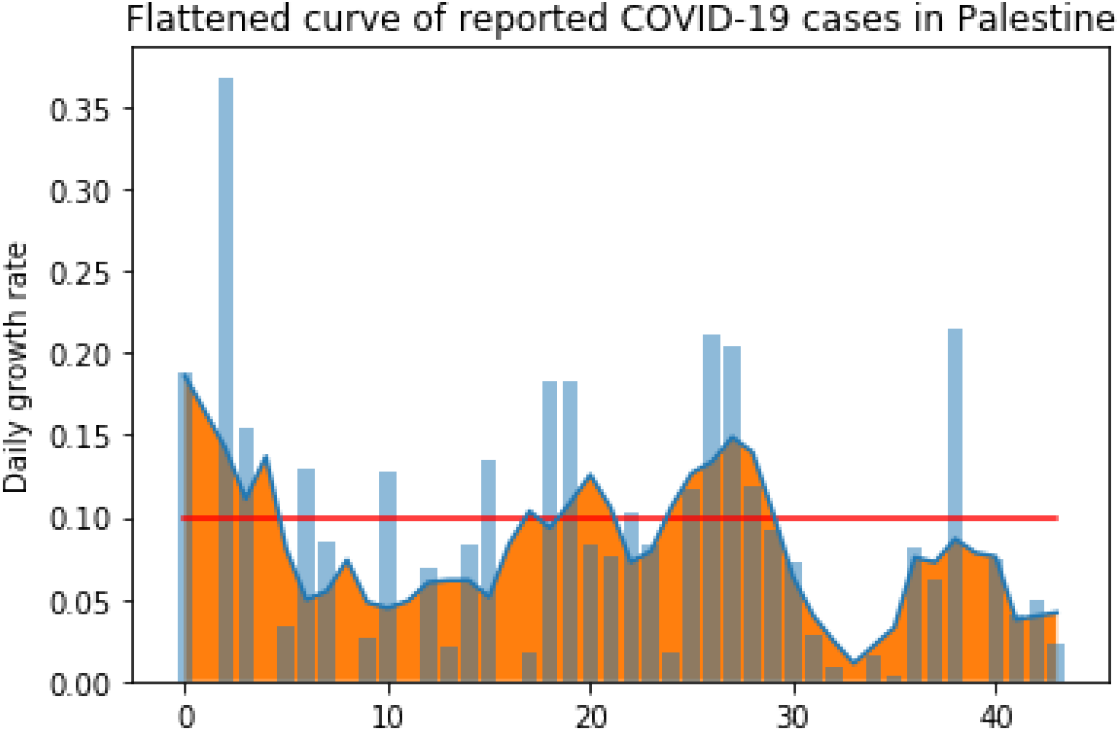
Flattened curve of daily reported infected cases in Palestine from March 5 until 22^*th*^ April 2020.

Patients who die on a given day *j* were infected much earlier, so the mortality rate denominator should be the total number of patients infected at the same time as those who died [12]. The mortality rate is defined by 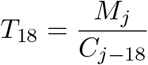 where *M*_*j*_ is the total number of deaths at time *j*. The median mortality rate during this period is equal to 0.03. The progression of the number of cases reported over the last eighteen days *P*_18_ is known in each country. It depends on the rate of containment and its effectiveness: 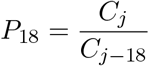.

Figure 3 shows the progression rate of COVID-19 in Palestine. The median of *P*_18_ is equal to 3.95 and it is used to compute the estimated number of reported and unreported cases [1]. We estimate the number of cases in Palestine as of April 22^*th*^, 2020 by modulating the estimated mortality rate. In Figure 4 we take different values of *T*: from 0.5 to 2%, where *P*_18_ = 3.38 with an account delays of 18 days between the onset of symptoms (or asymptomatic) and death. The more unfavorable case is 2 026 infected (unreported and unreported cases) and the optimistic simulation predicts 506 infected cases on 22 April.

**Figure 3:**
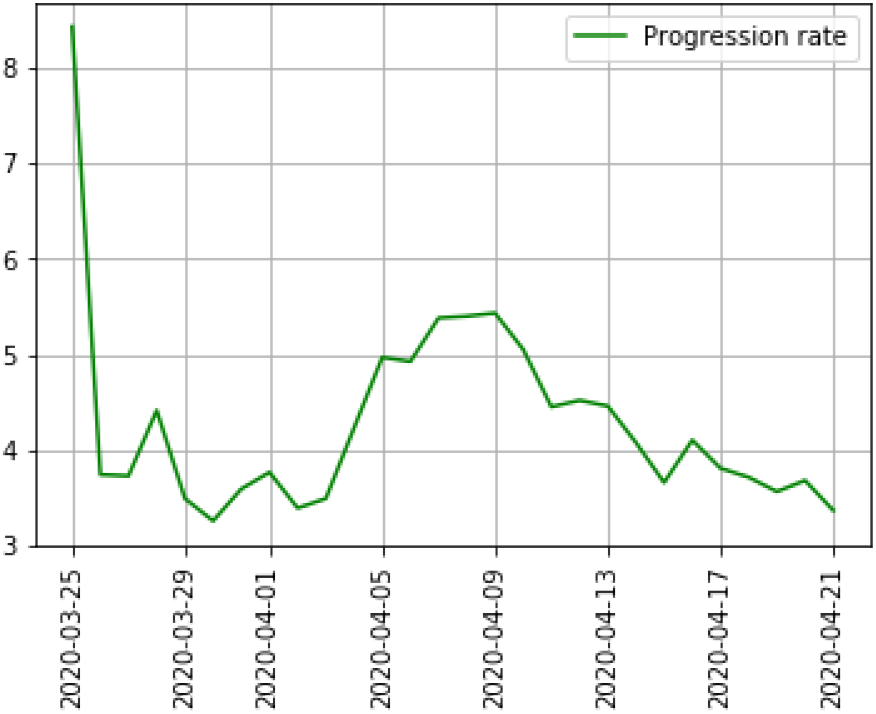
Progression rate *P*_18_ of COVID-19 in Palestine. The progression rate *P*_18_ is equal to 8.43 on March 25 and decreases to 3.38 on April 22 2020.

**Figure 4:**
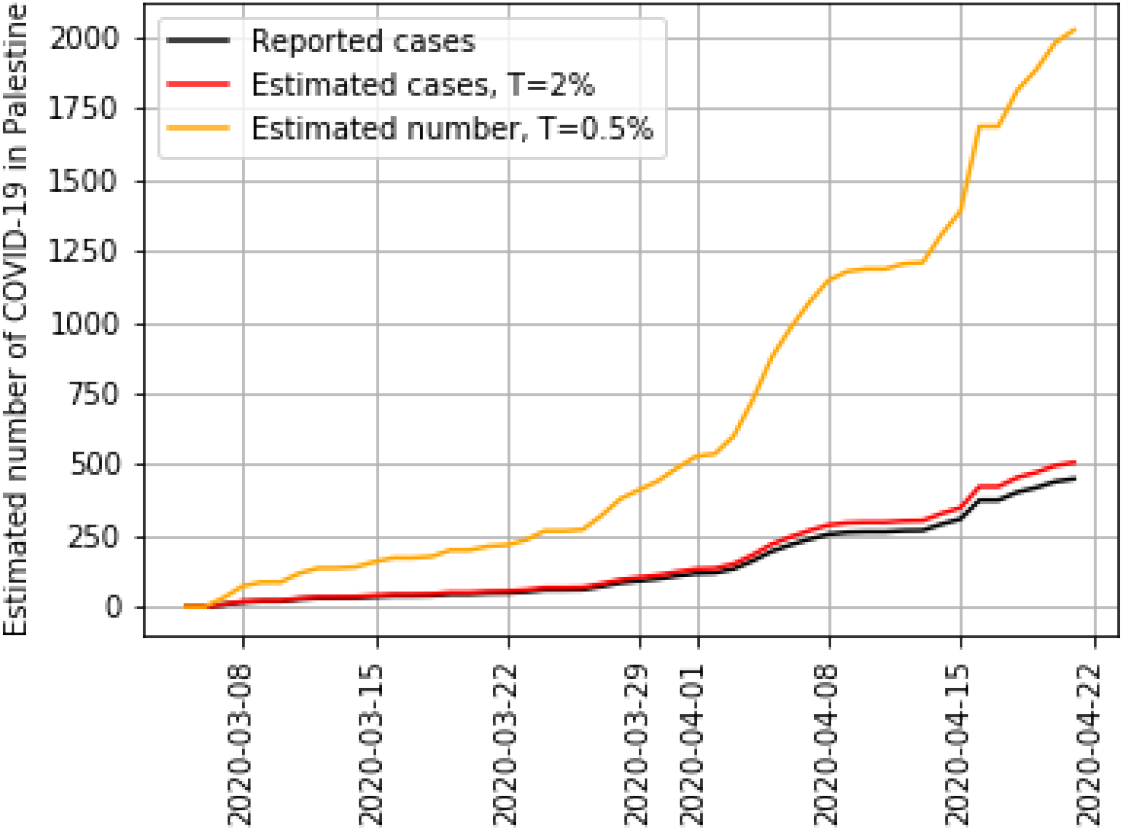
Estimated infected cases of COVID-19 in Palestine. Simulations using the average time, from first symptom to death, 18 days. The worse case is 2 026 and the optimistic status on April 22^*th*^ is 506 infected cases.

On the other hand, we scale the number of deaths per 1,000 population due to COVID-19 related to the number of registered infected people per million population. To measure a death ratio, a delay of 18 days is used between the mortality number and the infected number. This delay is justified by the days between infection and death, which is about 17 to 19 days [3]. The mortality ratio per thousand to infected cases per million is shown in Table 1. eatures are detailed in Figure 13. The relationships between deaths a given day *j* and the number of cases 18 days before *j* − 18 is linear for several countries.

**Table 1:** Ratio of deaths per thousand to infected cases per million, multiplied by 10^6^

| Jordan | Palestine | Egypt | Lebanon | Iran | Israel | Syria | Iraq | Saudi Arabia | Turkey |
| --- | --- | --- | --- | --- | --- | --- | --- | --- | --- |
| 16 | 7 | 76 | 31 | 63 | 13 | 71 | 52 | 9 | 24 |

**Figure 13:**
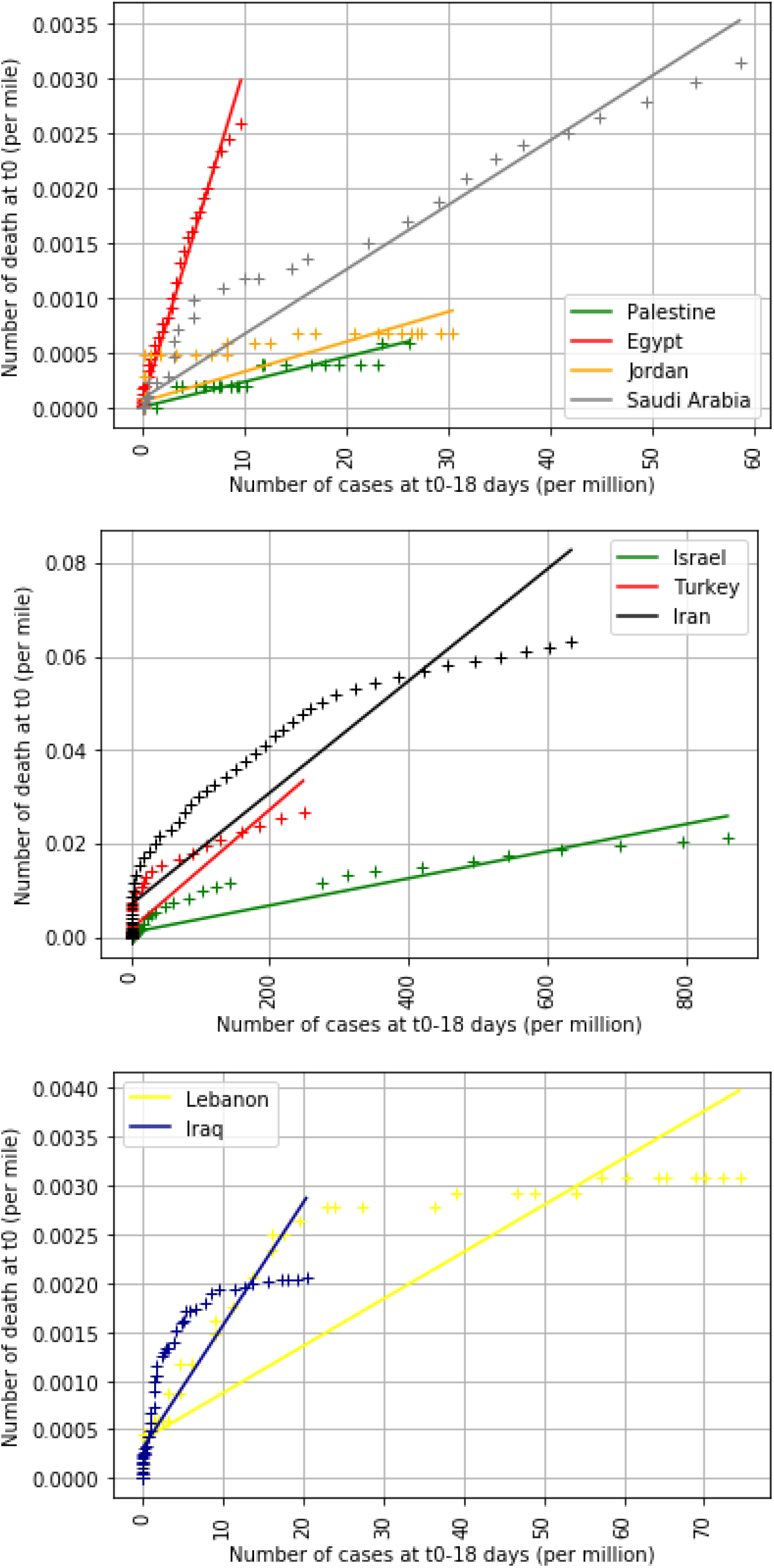
Number of deaths at *t*_0_ as a function of the number of cases eighteen days earlier (*t*_0_-18 days). The data are divided into three groups according to the number of deaths (depending on whether the number of deaths is lower in the country)

Compared to its neighbors, Palestine appears to be doing rather well, with a ratio of deaths per thousand to infected cases per million equal to 7 10^−6^. The actual number of cases as of April 22 is 466. This number per million persons puts Palestine, Egypt, Jordan, Syria, Iraq, and Lebanon in the same group (2.4 for Syria and 99.2 for Lebanon per million persons, Palestine has an infection rate equal to 88.4 per million). The second block of countries is Israel, Turkey, Iran, and Saudi Arabia (334.1 for Saudi Arabia and 1610 for Israel per million people). The rate of deaths provides two groups: the first with Israel, Turkey, Iran (0.021, 0.027, 0.063 per thousand people). And a second one with the rest of the 7 other countries.

## 3 Methods

Assume that infected individuals were not infectious during the incubation period [13]. Assume population growth rate and death rate are zero. Assume people exhibit consistent behaviors before and during the epidemic [14]. Assume no quarantine or other mitigation intervention is implemented. All these assumptions directly influence the model, which is dependent on several parameters. See [10] for a more precise idea of the evolution of these assumptions. The purpose of the model is to predict forward in time the future number of cases in a time-line of the epidemic from early reported case data. A typical SEIR (susceptible, exposed, infectious, removed) model is done in [4]. It can be described as a system of differential equations:

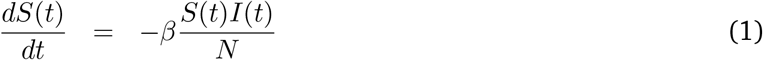

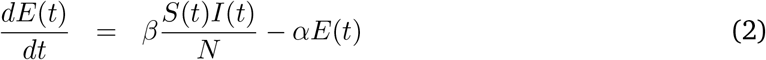

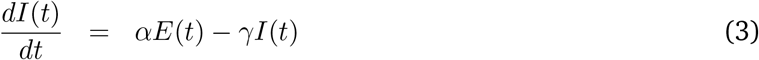

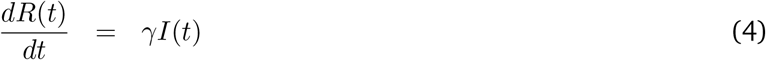

where, *t*_0_ is the beginning date of the epidemic, *t* ≥ *t*_0_ is time in days, *S*(*t*) is the number of susceptible at time *t, E*(*t*) is the number of exposed at time *t, I*(*t*) is the number of infectious at time *t, R*(*t*) is the number of removed, which includes the number of recovered and dead at time *t, N* (*t*) is the population at time *t* and *N* (*t*) = *S*(*t*) + *E*(*t*) + *I*(*t*) + *R*(*t*). Between *S* and *I*, the transition rate is 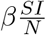, where *β* is the average number of contacts per person per time, multiplied by the probability of disease transmission in a contact between a susceptible and an infectious subject, and *I*/*N* is the fraction of contact occurrences that involve an infectious individual. The transition rates are *α* and *γ*.

We modify the model to estimate the number of unreported infected and asymptomatic cases out of the total number of cases. We propose to use a mathematical model, which recovers from data of reported cases, the number of unreported cases for the COVID-19 epidemic in Palestine. A sophisticated SEIR model is developed in [11], but we choose to folow the approach of [5] and the model in [6] to have a starting point of view of the epidemic. It produces a result closer to the realistic basic reproductive number ℛ_0_.

Incorporating revisions to the SEIR model of Figure 5 produces Figure 6. The parameters are listed in Table 2. The model contains a symptom class with both reported and unreported cases. The SEIR is given by the following system of ordinary differential equations:

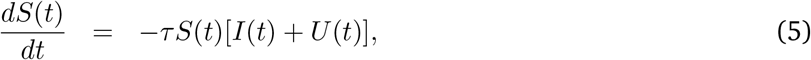

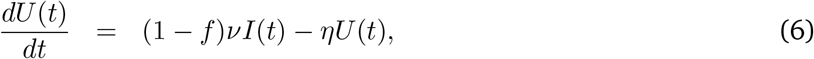

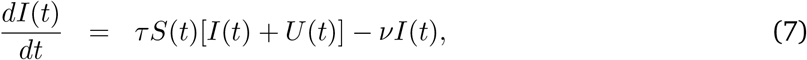

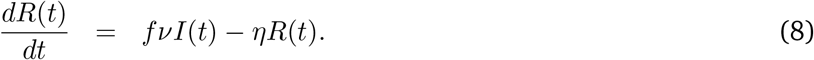

**Table 2:** Parameters and initial conditions of the model

| Symbol | Interpretation | Method |
| --- | --- | --- |
| $t_0$ | Time at which the epidemic started | fitted |
| $S_0$ | Number of susceptible at time $t_0$ | **fixed** |
| $I_0$ | Number of asymptomatic infectious at time $t_0$ | fitted |
| $E_0$ | Number of unreported symptomatic infectious at time $t_0$ | fitted |
| $\tau$ | Transmission rate | fitted |
| $1/\nu$ | Average time during which asymptomatic infectious are asymptomatic | **fixed** |
| $f$ | Fraction of asymptomatic infectious that become reported symptomatic infectious | **fixed** |
| $f\nu$ | Rate at which asymptomatic infectious become reported symptomatic | fitted |
| $(1-f)\nu$ | Rate at which asymptomatic infectious become unreported symptomatic | fitted |
| $1/\eta$ | Average time symptomatic infectious have symptoms | **fixed** |

**Figure 5:**
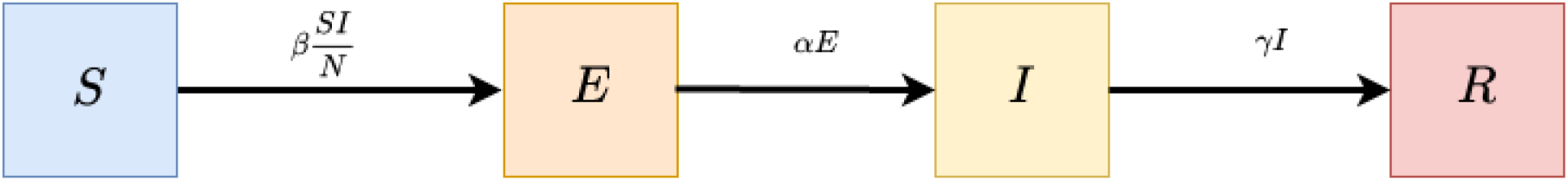
Diagram flux.

**Figure 6:**
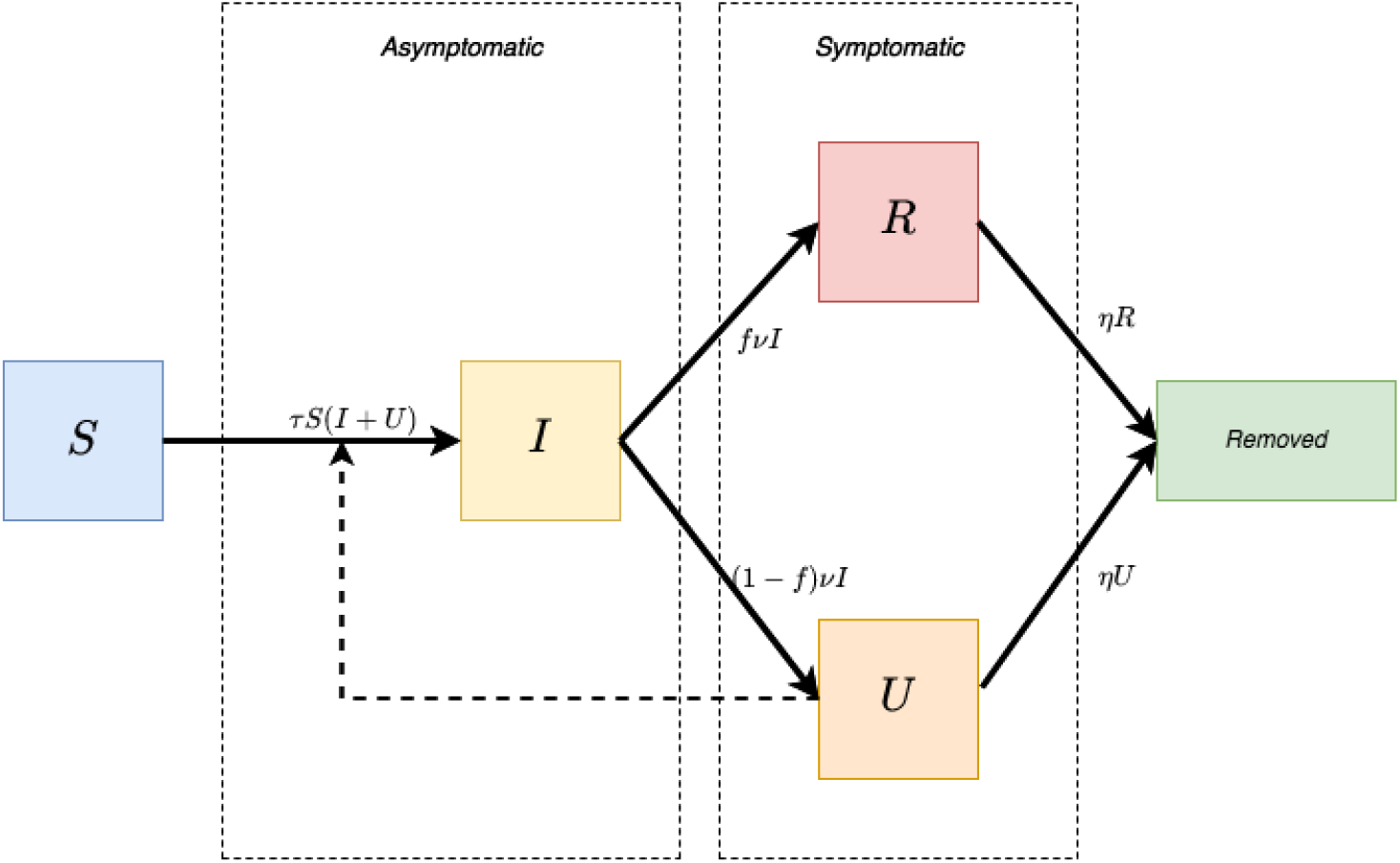
Diagram flux.

Here, *S*(*t*) is the number of individuals susceptible to infection at time *t, I*(*t*) is the number of asymptomatic infectious individuals at time *t, R*(*t*) is the number of reported symptomatic infectious individuals (i.e. symptomatic infectious with severe symptoms) at time *t*, and *U* (*t*) is the number of unreported symptomatic infectious individuals (i.e. symptomatic infectious with mild symptoms) at time *t*. This system is supplemented by initial data

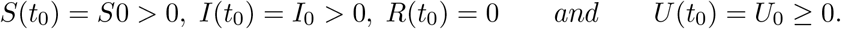

We assume that the removal rate *ν* is the sum of the removal rate of reported symptomatic infectious individuals, and of the removal rate of unreported symptomatic infectious individuals due to all other causes, such as mild symptom, or other reasons. The cumulative number of reported symptomatic infectious cases at time *t* is denoted by *CR*(*t*). We assume that *CR*(*t*) has the following special form:

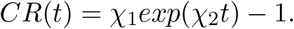

We obtain the model starting time of the epidemic *t*_0_:

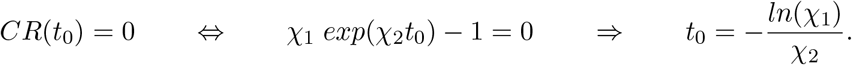

We fix *S*_0_ to 5 077 760, which corresponds to the total population of Palestine. We assume that the variation in *S*(*t*) is small during the period considered, and we fix *ν, η, f*. We can estimate the parameter *τ* and the initial conditions *U*_0_ and *I*_0_ from the cumulative reported cases *CR*(*t*). We then construct numerical simulations and compare them with data. We obtain *I*(*t*) = *I*_0_ *exp*(*χ*_2_(*t* − *t*_0_)) and 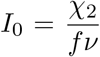. We must have *U* (*t*) = *U exp*(*χ* (*t* − *t*)). So, by substituting these expressions into previous identities, we obtain: *χ*_2_ *I*_0_ = *τ S*_0_ (*I*_0_ + *U*_0_) − *νI*_0_, *χ*_2_ *U*_0_ = (1 − *f*)*ν I*_0_ − *ηU*,

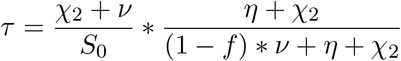

and 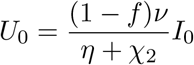. We fix *τ* in such a way that the value *χ*_*2*_ becomes the dominant eigenvalue of

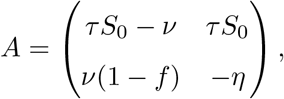

and (*I*_0_, *U*_0_) is the positive eigenvector associated with this dominant eigenvalue *χ*_2_. Thus, we apply implicitly the Perron–Frobenius theorem. Moreover, the exponentially growing solution (*I*(*t*), *U* (*t*)) that we consider (which is starting very close to (0, 0)) follows the direction of the positive eigenvector associated with the dominant eigenvalue *χ*_2_. The basic reproductive number becomes:

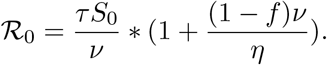

## 4 Comparison of models with the data

The first infected case in Palestine is documented on March 5, 2020. It is used as the first day of the forecast for Palestine. Fitting Palestinian data from April 15 to April 22 enables to compute *CR*(*t*) = *χ*_1_*exp*(*χ*_2_*t*) − 1. We find *χ*_1_ = 30.91 and *χ*_2_ = 3.16. This model shows that the starting of the epidemic is February 15, 2020 (the value of *t*_0_ = −59). As long as the number of reported cases follows *CR*(*t*), we can predict the future values of reported cases in Figure 11. For *χ*_1_ = 30.91, *χ*_2_ = 0.06 and *χ*_3_ = 1, we obtain prediction in Table 3. Thus, the exponential formula overestimates the number of infected cases after 4 months (see Figure 12). From now on, we fix the fraction *f* = 0.6 of asymptomatic that become reported symptomatic infectious. The average time during which asymptomatic infectious cases are symptomatic is equal to 1/*ν* = 1/7. The average time symptomatic infectious have symptoms is equal to *η* = 1/7. The values 1/*η* = 7 days and 1/*ν* = 7 days are taken from information concerning earlier corona viruses, and are used now by medical authorities [9].

**Table 3:** Data for Palestine - accumulated reported cases: estimated CR(t)

| 23 April | 24 April | 25 April | Mai 1st | 5 May | June 1st |
| --- | --- | --- | --- | --- | --- |
| 543 | 576 | 611 | 869 | 1099 | 5344 |

**Figure 11:**
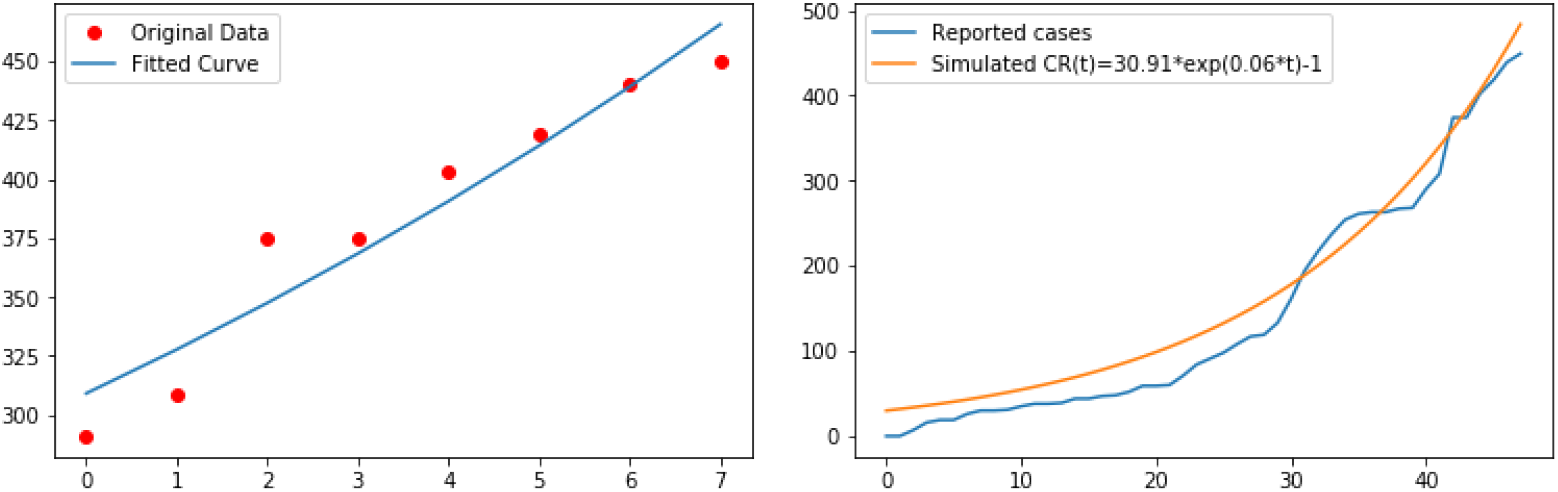
In the left side, fitted reported cases *CR*(*t*) to Palestinian data from April 15 to April 22, 2020. In the right side, fitted reported cases *CR*(*t*) = 30.91 exp(0.06 *t*) − 1 to Palestinian data from March 5 to April 22, 2020.

**Figure 12:**
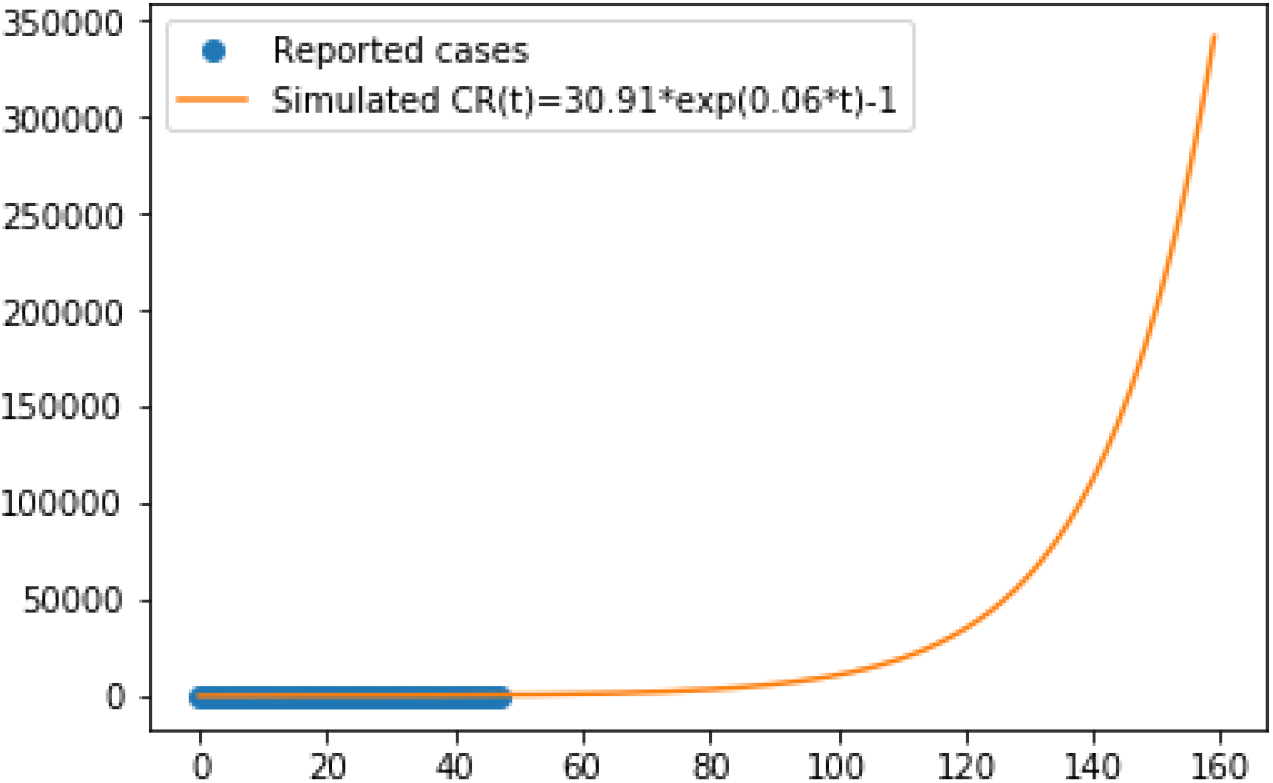
In this figure, we use *CR*(*t*) = 30.91 exp(0.06*t*) −1, which corresponds to the estimated infected cases with respect to data for Palestine in Table 4.

For *N* a total population equal to 5 077 760, we suppose that *S*_0_ = *N*/100 is the susceptible population to be infected by COVID-19. Figure 7 and Figure 8 show that the range between the most naive model and the slightly more elaborate one is between 113 000 and 11 014 infected cases. But both converge towards the first peak of the pandemic at the end of May 2020. The results of this model, if correct, show very good management of the epidemic regarding the number of deaths compared to its neighboring countries.

**Figure 7:**
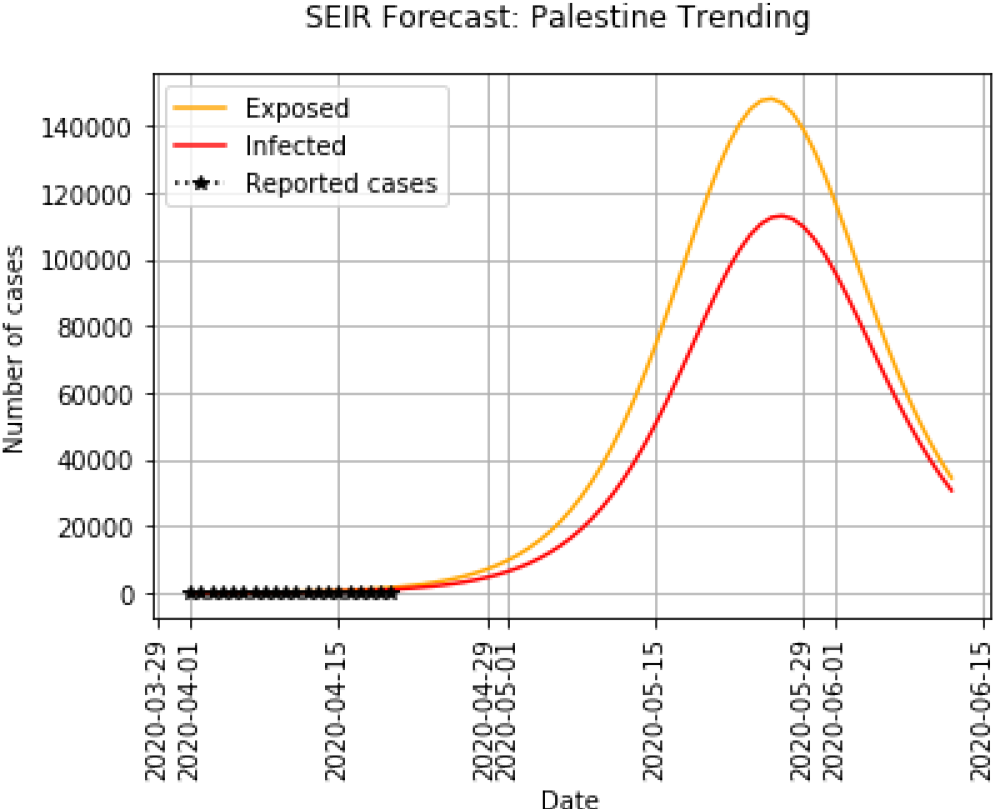
SEIR with *α* = 0.74, *β* = 1.38, *γ* = 0.96, a maximum number of infected cases 113 171 obtained on 2020-05-27.

**Figure 8:**
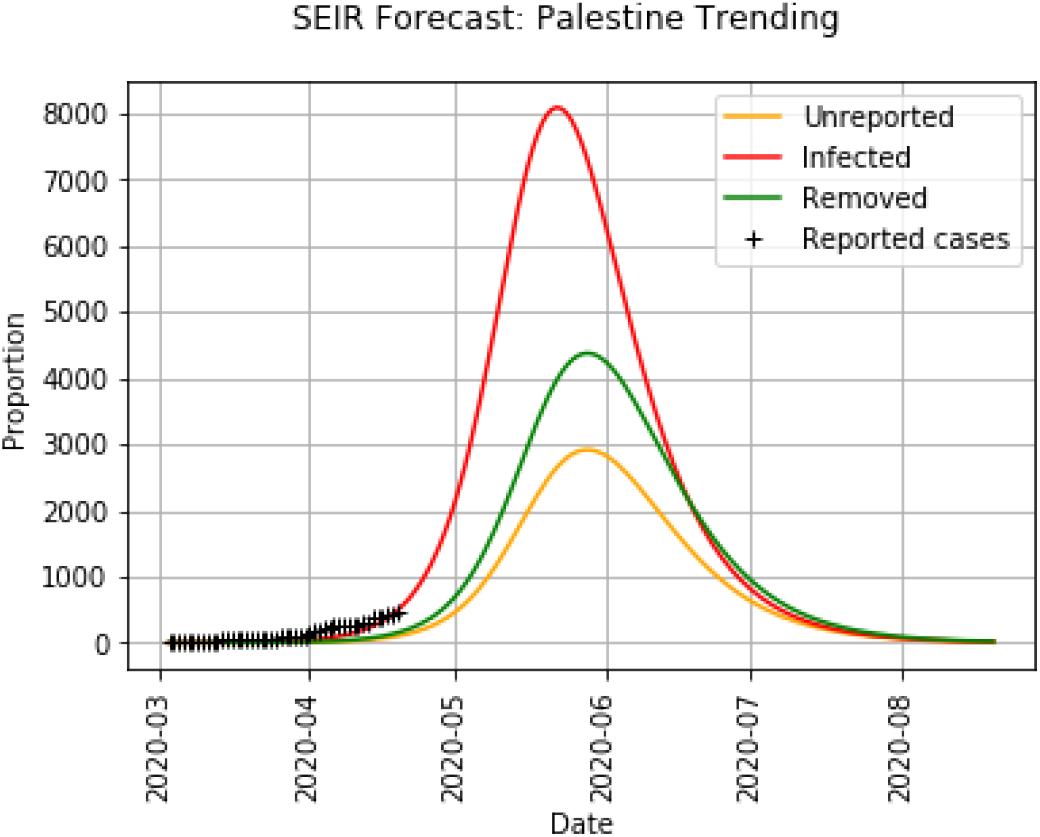
SEIR with *f* = 0.6, *η* = 1/7, *ν* = 1/7, *τ* = 4.55 10^−6^ and ℛ_0_ = 1.54. 8 095 reported cases and 2 919 unreported cases, with a maximum cases on 2020-05-22.

For these values of parameters, ℛ_0_ = 1.54. In the literature, the value of ℛ_0_ varies from 1.4 to 3.9 [8]. Our simulations in Figure 9 give amounts that belong to this range. We estimated when the effective daily reproduction ratio has fallen below 1. When reported symptomatic individuals *R*(*t*) are infectious for an average period of 1/*η* = 7 days, the transmission rate *τ* = 4.55 10^−6^ and the average period of infectiousness of asymptomatic infectious individuals is 1/*ν* = 2 days: if *f* = 0.7 then ℛ_0_ = 0.95 and if *f* = 0.8 then ℛ_0_ = 0.79. The codes for the simulation are available on GitHub^2^.

**Figure 9:**
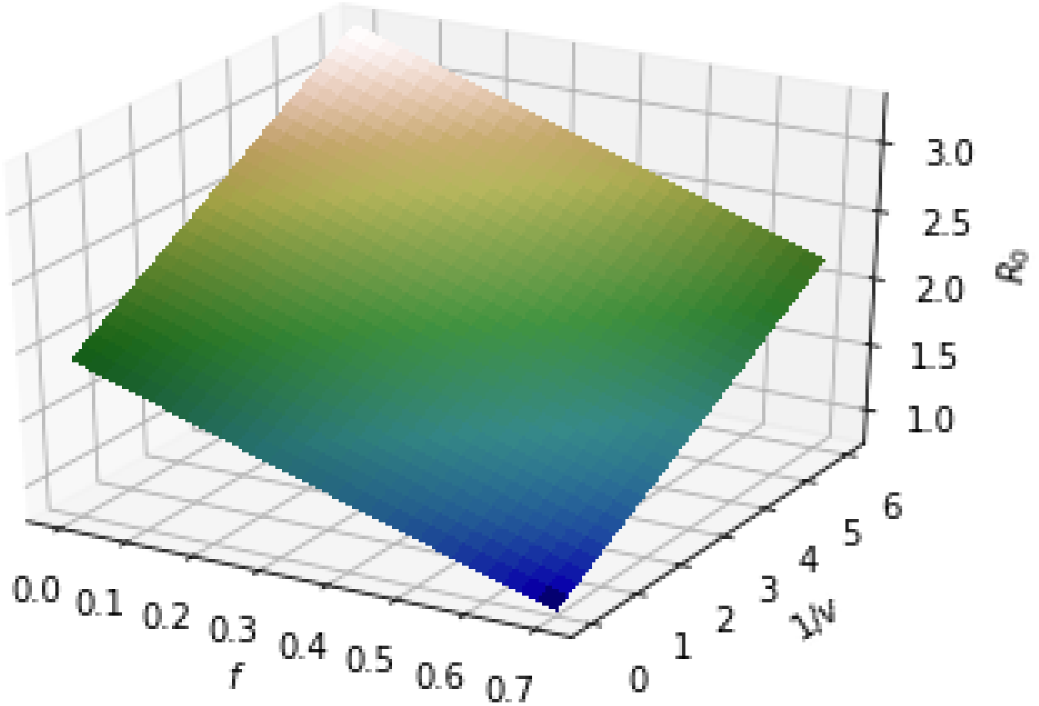
In this figure, we use 1/*η* = 7 days, and we plot the basic reproductive number ℛ_0_ as a function of *f* and 1/*ν* using *τ* = 4.55 10^−6^, which corresponds to the data for Palestine in Table 4.

**Figure 10:**
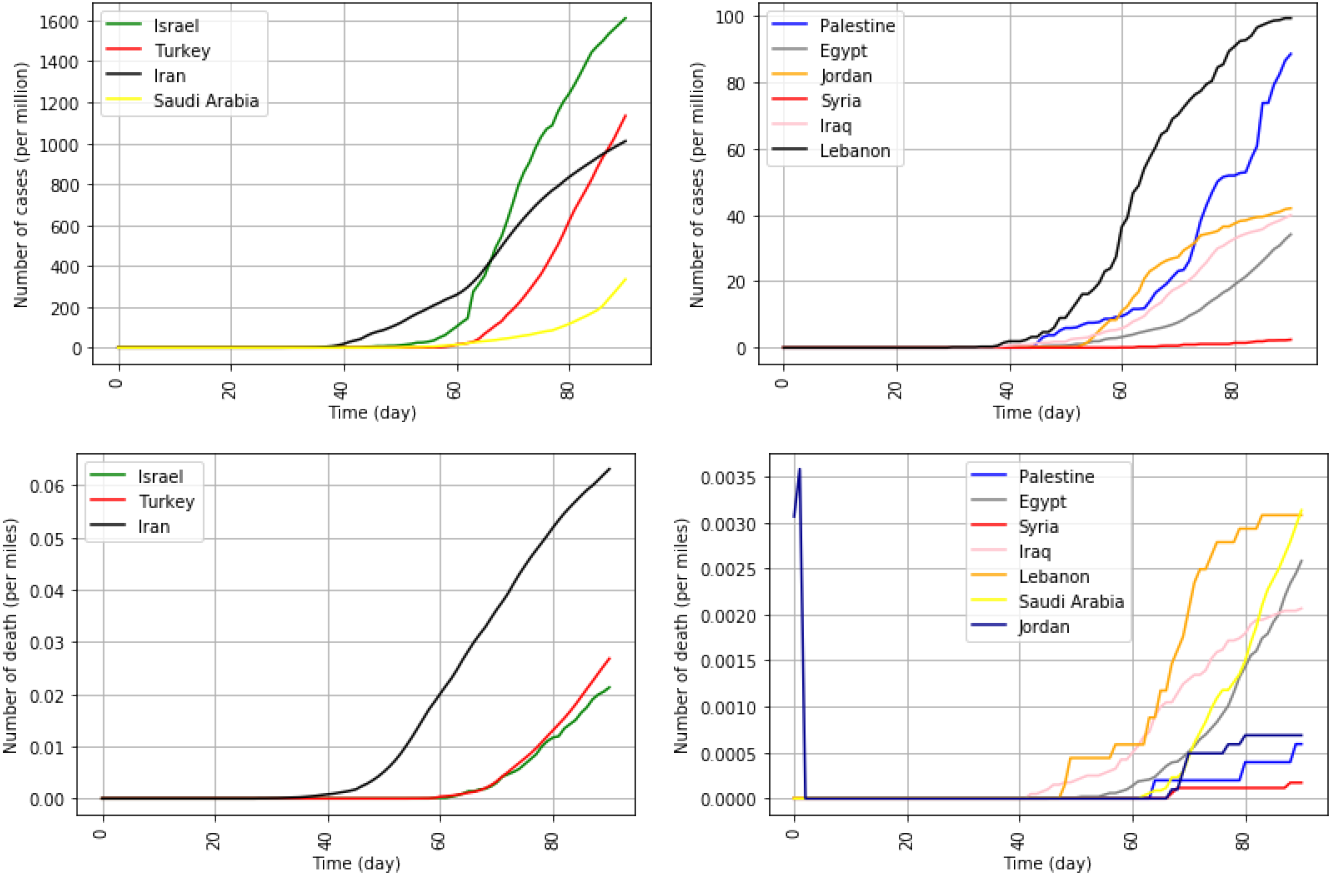
Number of infected cases and deaths. The data are divided into different groups. These number are normalized per million for reported infected cases or per miles for death cases.

## 5 Discussion

An epidemic outbreak of a new human coronavirus COVID-19 will occur in Palestine. The unreported cases and the disease transmission rate are useful information. We estimate an actual number of infected cases in Palestine based on the 18-day effect from infection to death. We find that the number of cases in April 22 varies between 506 and 2 026 infected cases. To recover these effect of unreported cases, we applied a method developed in [7] to predict the evolution of a COVID-19 epidemic in Palestine, based on reported case data.

On April 22, the number of cases is 466 infected, 4 dead, and a total population of 5 077 760. The number of infected cases per million persons puts Palestine, Egypt, Jordan, Syria, Iraq, and Lebanon in the same package (between 2.4 for Syria and 99.2 for Lebanon per million people, Palestine has an infection rate equal to 88.4 per million people). The second group of countries is Israel, Turkey, Iran, and Saudi Arabia (334.1 for Saudi Arabia and 1610 for Israel per million people). The number of death cases also gives two blocks: one with Israel, Turkey, Iran (21, 27, 63 per million people). And a second one with the rest of the 7 other countries (0.6 per million for Palestine, 0.7 per million for Jordan and 3 per million for Lebanon). Figure 13 shows a good management of the epidemic by Palestine, especially on the number of deaths, compared to countries neighboring Palestine.

In the case of Palestine, the peak of the epidemic occurred approximately May 22. Accordingly to the SEIR model with asymptomatic and symptomatic compartments, the total number of cases reaches a maximum of approximately 11 014 cases near the turning point May 22. The worst scenario predicts 113 171 infected cases (with a classic SEIR approach). The crest of the pandemic is from 22 to 27 May 2020. The reproductive number ℛ_0_ is equal to 1.54 for a fixed fraction of *f* = 0.6 of symptomatic cases that are reported and for a removal rate of *ν* = 7. The epidemic can be contained in Palestine, provided that quality of care and social distancing measures are in place. We could then hope to see the reproductive number of COVID-19 in Palestine in Figure 9 dropping to an amount less than 1.

To estimate the reproductive number ℛ_0_, we initially get the time-dependent function of reported cases *CR*(*t*). With the SEIR method used, the reproductive number is very sensitive to the data set: Formula for *CR*(*t*) is descriptive for the reported case data for Palestine, and ℛ_0_ is still greater than 1. To reduce ℛ_0_ to less than 1, public measures should be taken, such as isolation, quarantine, and public closings. These measures exacerbate the spread of this disease and mitigate the final size of the epidemic [11]. The model used in this paper incorporates social distancing measures through the transmission rate *τ* = 4.55 10^−6^. The consequences of late public interventions may have severe consequences for the epidemic outcome. Changes to this rate (over time) would be advisable when the epidemics will peak.

## Data Availability

All data are available in https://www.worldometers.info/coronavirus/ and https://corona.ps/details

## Acknowledgement

The author would like to thank Pr. Ahmed Abbes for his valuable feedback, comments and rewarding discussions.

## Appendix

In Figure 9, we graph ℛ_0_ as a function of *f* and 1/*ν*, to illustrate the importance of these values in the evolution of the epidemic. The accuracy of these values depend on the input of medical and biological epidemiologists. We find that minimum value of ℛ_0_ is equal to 1.18 (*f* = 0.7 and 1/*ν* = 3), the maximum is equal to 3.3 (*f* = 0.1 and 1/*ν* = 8) and the median is equal to 2.24.

## Footnotes

1 Data is available on https://www.worldometers.info/coronavirus/ and at https://corona.ps/details

2 https://github.com/inestej/PalestinianCOVID19

